# Integrated analyses of single-cell RNA-seq public data reveal the gene regulatory network landscape of respiratory epithelial and peripheral immune cells in COVID-19 patients

**DOI:** 10.1101/2023.03.09.23287043

**Authors:** Lin Zhang, Hafumi Nishi, Kengo Kinoshita

## Abstract

**Introduction:** Infection with SARS-CoV-2 leads to coronavirus disease 2019 (COVID-19), which can result in acute respiratory distress syndrome and multiple organ failure. However, its comprehensive influence on pathological immune responses in the respiratory epithelium and peripheral immune cells is not yet fully understood.

**Methods:** In this study, we integrated multiple public scRNA-seq datasets of nasopharyngeal swab and peripheral blood results to investigate the gene regulatory networks (GRNs) of healthy individuals and COVID-19 patients with mild/moderate and severe disease, respectively. Similar and dissimilar regulons were identified within or between epithelial and immune cells during COVID-19 severity progression. The relative transcription factors (TFs) and their targets were used to construct GRNs among different infection sites and conditions.

**Results:** Between respiratory epithelial and peripheral immune cells, different TFs tended to be used to regulate the activity of a cell between healthy individuals and COVID-19 patients, although they had some TFs in common. For example, XBP1, FOS, STAT1, and STAT2 were activated in both the epithelial and immune cells of virus-infected individuals. In contrast, severe COVID-19 cases exhibited activation of CEBPD in peripheral immune cells, while CEBPB was exclusively activated in respiratory epithelial cells. Moreover, in patients with severe COVID-19, CEBPD upregulated S100A8 and S100A9 in CD14 and CD16 monocytes, while S100A9 genes were co-upregulated by different regulators (SPEDEF and ELF3) in goblet and squamous cells. The cell-cell communication analysis suggested that epidermal growth factor receptor signaling among epithelial cells contributes to mild/moderate disease, and chemokine signaling among immune cells contributes to severe disease.

**Conclusions:** This study identified cell type- and condition-specific regulons in a wide range of cell types from the initial infection site to the peripheral blood, and clarified the diverse mechanisms of maladaptive responses to SARS-CoV-2 infection.

## 1 Introduction

Severe acute respiratory syndrome coronavirus 2 (SARS-CoV-2) infection causes a contagious disease known as Coronavirus Disease 2019 (COVID-19), which spread quickly globally and resulted in the COVID-19 pandemic. According to the World Health Organization (WHO), there had been more than 619 million cases worldwide (including over 6.5 million deaths) by the beginning of October 2022. Although the vast majority of infected individuals have asymptomatic, moderate, or mild symptoms, a proportion of cases require hospitalization and intensive care, or even progress to death (1–4). SARS-CoV-2 enters epithelial cells, assembles its structures and nucleocapsids, is released, and subsequently stimulates immune cells (such as macrophages and dendritic cells) by inducing inflammatory factors. Finally, its antigen is presented via histocompatibility complexes I and II (MHC I and II) to activate humoral and cellular immunities that are mediated by B and T cells to induce the production of cytokines and antibodies (5–10). The severity of inflammation can lead to cytokine storms in some COVID-19 patients (11–14).

COVID-19 affects patients differently, and distinct features have been noted. For example, immunological signatures are altered during severe infection, and levels of a wide range of pro-inflammatory cytokines (such as S100A8/A9, interleukin 1 beta, interleukin 6 (IL-6), IL-8, CXCL10, and tumor necrosis factor alpha (TNFα)) are dramatically increased (15–18). Compared to severe disease, the substantial expression of genes associated with interferon (IFN) responses (type I in particular) has been observed in cells (such as epithelia) in mild or moderate COVID-19 disease (19–21). Additionally, patients with severe COVID-19 show activation of neutrophils (22,23) and lymphocyte exhaustion (24,25). Given the distinct antiviral immunity among cell types during the progression of SARS-CoV-2 infection, various therapeutic strategies have been developed to improve COVID-19 treatment (13,26). For example, targeting cytokine storms improves outcomes and reduces mortality in elderly patients with COVID-19 (27). In this respect, Tocilizumab, an IL-6 pathway inhibitor, improved the clinical manifestations in 21 patients with severe and critical COVID-19 (28).

Multiple studies have been conducted to date to investigate alterations associated with immune responses, with the aim of providing deeper insights into the roles of the nasal, upper, and lower airway tissues and peripheral blood (21,29–32). A large-scale single-cell transcriptome atlas of the lungs and peripheral blood of COVID-19 patients has also been compiled (33). However, a detailed analysis of the gene regulatory changes in both respiratory epithelial and peripheral immune cells during progression to severe COVID-19 is required to completely understand aberrant and protective immune responses to SARS-CoV-2 infection.

Therefore, in this study, we integrated single-cell RNA sequencing data from nasopharyngeal swabs and peripheral blood mononuclear cells (PBMCs) to capture the immune response at the site of infection (epithelial cells) and of the peripheral immune system. We found that epithelial cells (e.g., squamous and goblet cells) and immune cells (e.g., CD14 and CD16 monocyte cells) exhibit substantial phenotypic differences after SARS-CoV-2 infection. The transcription factor regulatory network construction underlies heterogeneous immune responses during progression to severe COVID-19 among cell types from different infection sites. Furthermore, we demonstrated the important role of some inflammatory genes (such as S100A8 and S100A9) in the pathogenesis of COVID-19 and found that regulators of these critical genes can be unique to cell types and conditions. A cell-cell communication analysis suggested that epidermal growth factor receptor (EGFR) signaling in epithelial cells may contribute to mild/moderate COVID-19. Collectively, our work reveals and clarifies the mechanisms involved in maladaptive responses to SARS-CoV-2 infection and provides a rich resource for predicting, preventing, and treating SARS-CoV-2 infection in respiratory epithelial cells and peripheral immune cells.

## 2 Materials and Methods

### 2.1 Data collection

Single-cell RNA sequencing (scRNA-seq) data from nasopharyngeal swabs and PBMCs were collected (21,31). The nasal scRNA-seq data are publicly available for exploration and download via the single-cell portal (https://singlecell.broadinstitute.org/single_cell/study/SCP1289/), and the PBMCs data are available for viewing and downloading from the COVID-19 Cell Atlas (https://www.covid19cellatlas.org/#wilk20) hosted by the Wellcome Sanger Institute.

Biological samples of nasopharyngeal swabs were collected from the University of Mississippi Medical Center between April and September 2020, and eligible participants for blood samples were recruited into the Stanford University ICU Biobank study between March 2020 and April 2020. With respect to the nasal epithelial data, eight individuals were removed from our study based on the following criteria: (1) healthy individuals with a recent history of COVID-19 and (2) individuals who needed intensive care units but without a recent history of COVID-19. In addition, because of the small numbers of cells collected from mast cells (6 cells), plasmacytoid DCs (11 cells), and enteroendocrine cells (1), these cell types were excluded from our analyses. A total of 15 healthy participants and 35 patients diagnosed with COVID-19 were ultimately studied. According to the COVID-19 severity stratification of the World Health Organization (WHO) guidelines, these 35 patients were further divided into two groups: those with mild/moderate disease (14 patients) and those with severe disease (21 patients). WHO scoring system for healthy, mild/moderate, and severe cells were represented by Control_WHO_0, COVID19_WHO_1–5, and COVID19_WHO_6–8. With respect to the PBMC scRNA-seq data, six healthy and seven severely ill individuals were studied.

Processed count matrices with embeddings were used only for the PBMCs. The cell types in studied scRNA-seq data were annotated using the original papers.

### 2.2 Single-cell RNA sequencing data processing

The Seurat package (version 4.0.4) (34) implemented in R (version 4.1.0) was used to explore the single-cell transcriptome data. The count matrices were normalized using Seurat NormalizeData. Specifically, the log-normalized method was used to normalize the total feature expressions per cell, multiply them by a scaling factor (10,000 by default), and further log-transform the results. Highly variable genes were then identified using the FindVariableFeatures function (3,000 top variable features were set). The percentage of mitochondrial genes was regressed out using the ScaleData function. The scaled data were passed to run a principal component analysis (PCA) dimensionality reduction algorithm. The FindNeighbors and FindClusters functions were then employed to cluster the cells, and a graph-based clustering algorithm that calculates the k-nearest neighbors and constructs a shared nearest neighbor graph, was applied to identify cell clusters. Nonlinear dimensionality reduction (RunUMAP function) and Uniform Manifold Approximation and Projection (UMAP) were then conducted to visualize the clustering results in two dimensions. To identify differentially expressed genes (DEGs) when comparing any two given groups, the FindMarkers function in Seurat was applied with the following configurations: test.use = “wilcox” (a Wilcoxon Rank Sum test), min.pct = 0.25, logfc.threshold = 0.25, and only.pos = FALSE. An additional adjusted P-value threshold of ≤ 0.05 was used for filtering DEGs.

### 2.3 Gene regulatory network analysis and visualization

To explore the regulatory landscape across cell types between healthy and COVID-19 patients, the SCENIC (single-cell regulatory Network Inference and Clustering, version 1.2.4) (35) tool was used. SCENIC is a set of tools that can infer transcription factors (TFs) and construct gene regulatory networks (GRNs) from scRNA-seq data. The required human RcisTarget database was downloaded from https://resources.aertslab.org/cistarget/. GENIE3 and RcisTarget in SCENIC were used to identify potential direct binding targets (called regulons) based on co-expression modules and a TF motif analysis. Here, the regulon represented one TF and its targets. Utilizing the AUCell algorithm, the activity of regulons in each individual cell were analyzed and evaluated by calculating the area under the recovery curve (AUC) score. To identify specific regulators of cell type-specifics and conditions (healthy, mild/moderate, and severe COVID-19), we calculated the average regulon activity by cell type in each condition and merged them to create an AUC score heatmap via the pheatmap package in R. The AUC score matrix of all regulons in each cell was submitted to the Seurat object to project the AUC (as well as TF expression) onto UMAP plots. In addition, in terms of each condition, the targets of the identified TFs in each cell type were filtered using the corresponding DEGs. Finally, GRNs for each cell type and condition, which comprised the observed TFs and their differentially expressed target genes, were constructed and displayed using Cytoscape software (version 3.9.1) (36).

### 2.4 Function and pathway enrichment analysis

Gene ontology (GO) and Kyoto Encyclopedia of Genes and Genomes (KEGG) enrichment analyses of gene sets of interest were performed using the clusterProfiler (version 4.0.5) (37) package in R. The GO enrichment analysis was conducted based on biological processes, and GO annotation data were provided by AnnotationHub. KEGG annotation data are available in the KEGG database (https://www.genome.jp/kegg/). An adjusted P-value ≤ 0.05 was considered significantly enriched.

### 2.5 Cell-cell communication analysis

The CellChat (version 1.5.0) (38) package in R was used to infer and analyze the cell-cell communication (CCC) among cell types. “Secreted Signaling” was set to explore intercellular communication networks, and the communication probability was then computed to infer the cellular communication network. To calculate the aggregated CCC network, the number of links was counted or the communication probability summarized. Collectively, based on gene expressions and prior knowledge of the interactions, all significant communications (ligand-receptor interactions) associated with signaling pathways from one cell type to other cell types were determined.

### 2.6 Whole blood bulk transcriptomic data analysis

Pre-processed whole-blood bulk transcriptomic data are publicly available for download at GEO (accession number GSE157103) (39). Transcript counts were normalized using the transcript per million (TPM) method. Samples were selected from a total of 126 samples according to the following criteria: (i) select COVID-19 infection samples and (ii) samples were removed if the sequential organ failure assessment (SOFA) scores were unknown. A final total of 56 samples were used in this study. The selected samples were then grouped based on the SOFA score and the Pearson’s correlations calculated between the TPM and SOFA scores.

## 3 Results

### 3.1 Single-cell characterization of nasopharyngeal swabs and PBMCs

To better understand and compare the host response to SARS-CoV-2 infection at the initial infection site and peripheral immune cells, we obtained single-cell RNA sequencing (scRNA-seq) data from nasopharyngeal swabs (21) and peripheral blood mononuclear cells (PBMCs) (31) under three different conditions: healthy individuals and COVID-19 infection patients with mild/moderate and severe disease. Metadata, such as cell type annotation and embedding of PBMC data, were mainly obtained from original studies. As mentioned in the Materials and Methods section, certain cells associated with nasal scRNA-seq data were removed in this study based on the criteria described therein. Data from nasopharyngeal swabs and PBMC were then processed using the same protocols, including those relating to data normalization, dimensionality reduction, and cell clustering, and the results were visualized on the UMAP plot. A total of 26,894 cells from the nasal mucosa and 44,721 cells from PBMCs were studied, comprising 15 and 13 cell types, respectively (Figure 1A, Figure S1, and Table S1). Of the cell types, SARS-CoV-2 induced CD14 monocyte expansion and NK cell loss, while the B and T cell abundances were similar between healthy and COVID-19 patients. In addition, ciliated and goblet cells from nasal epithelial cells and dendritic cells (DCs) from PBMCs exhibited the highest number of expressed genes.

**Figure 1.**
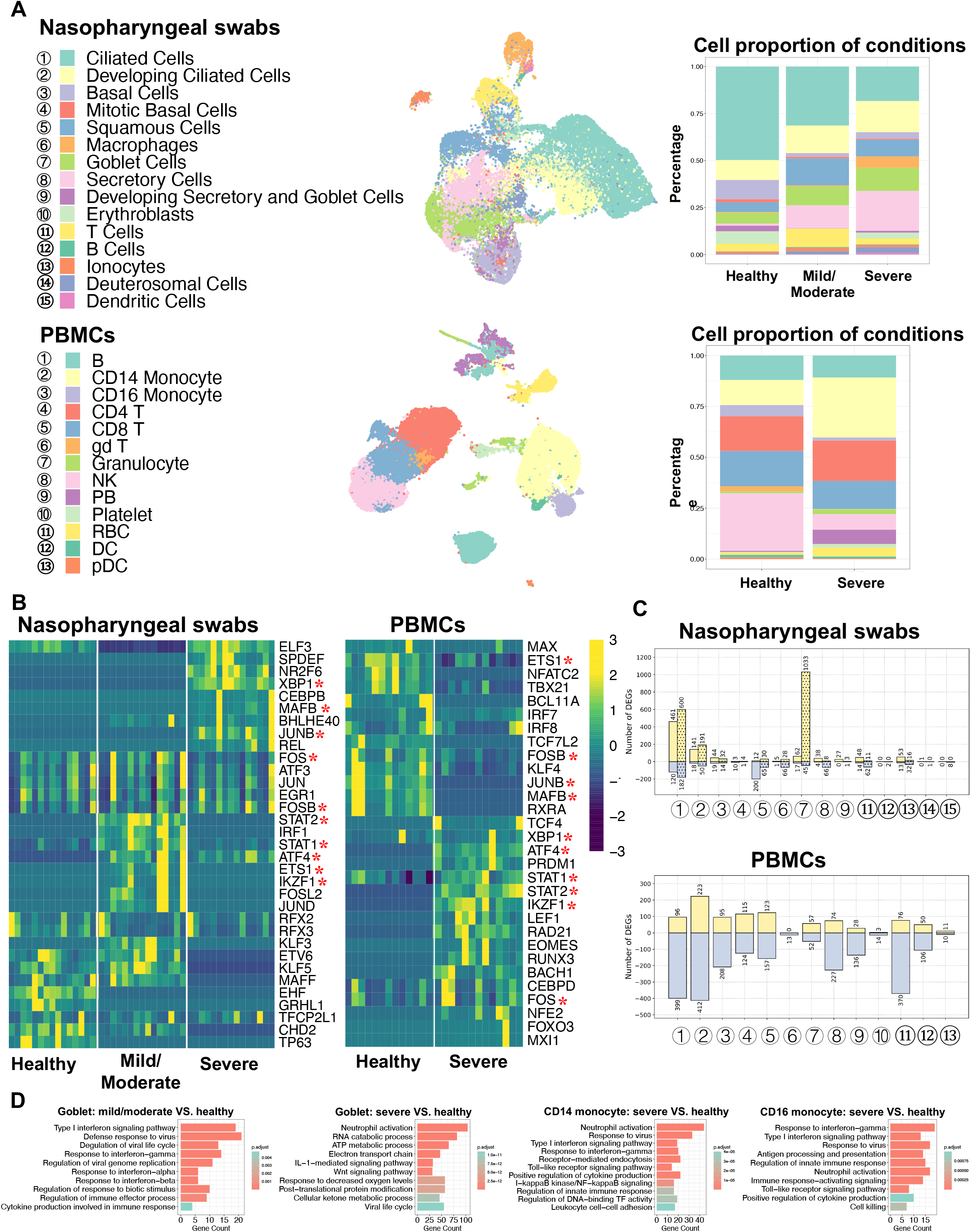
Characterization of nasopharyngeal swabs and PBMCs data. (A) Cell types in nasopharyngeal swabs and PBMCs data and their percentage proportion in patients with different COVID-19 severity. Middle: cell type visualization on UMAP plots. Right: cell proportion associated with disease severity (mild/moderate and severe) and healthy cells. (B) Heatmap of the area under the curve (AUC) scores of regulons estimated per cell type by SCENIC. Detected regulons are represented by their corresponding transcription factors in the right-hand columns. Columns represent AUC scores of cell types. For each condition, the column order is the same as the label order in (A). Asterisks behind the TFs represent commonly identified TFs between nasopharyngeal swabs and PBMCs. (C) Number of differentially expressed genes (DEGs) when comparing COVID-19 (mild/moderate, severe) and healthy cells. Yellow and light-blue colors represent upregulated and downregulated genes, respectively. The corresponding gene numbers are shown at the top and bottom of each bar. In the upper panel, bars without dots indicate identified DEGs by comparing mild/moderate disease to healthy cells, while bar with dots indicate DEGs when comparing severe disease and healthy cells. (D) GO (biological process) enrichment analysis to compare upregulated genes found in diseased and healthy cells.

### 3.2 Similarity and dissimilarity of regulons and pathways were identified in respiratory epithelial and peripheral immune cell types associated with COVID-19

To investigate the gene regulatory network (GRN) changes underlying COVID-19 manifestations, we conducted single-cell regulatory network inference and clustering (SCENIC) analyses (35).

Regulons, including transcription factors (TFs) and their direct target genes, were detected in each cell type. We then calculated the average area under the recovery curve (AUC) scores per cell type to estimate regulon activities. Using a SCENIC analysis, we identified potential TFs in terms of the cell type and three condition (healthy individuals and COVID-19 infections with mild/moderate and severe disease). By comparing nasopharyngeal swabs and PBMCs, we found that there were notable differences between the many identified regulons among conditions or infection sites (i.e., nasal or peripheral blood), while some were found to be shared, and XBP1, FOS, STAT1, and STAT2 were activated in both the epithelial and peripheral immune cells of virus-infected individuals (Figure 1B). In addition, when comparing the detected regulons across cell types, we found that some cell types were distinguished by different regulon combinations among the three conditions, but some were not (Figure S2A). Specifically, epithelial cells, such as ciliated cells and mitotic basal cells, unexpectedly shared common TFs, whereas distinct TFs were identified in other epithelia, such as basal cells, squamous cells, and goblet cells. For example, RFX2 and RFX3 showed high activity in ciliated cells regardless of disease severity, while XBP1, NR2F6, SPDEF, and ELF3 were preferentially activated in goblet cells in patients with severe COVID-19, and KLF5 and STAT2 were coactivated in patients with mild/moderate COVID-19. Furthermore, NR2F6, SPDEF, and ELF3 were found to be activated in squamous cells in patients with severe COVID-19, and this activity was also shared with goblet cells.

It is of note that we mainly focused on epithelial cells from nasal scRNA-seq data because we only obtained a very small number of immune cells, but we then used PBMC data to analyze immune cells. The PBMC data showed that most regulons were unique to cell types or patient condition (healthy and severe). For example, CEBPD and FOS were highly activated in both CD14 and CD16 monocyte cells in severe COVID-19 disease, whereas BACH1 exhibited particularly high activation in CD16 monocyte cells (Figure S3A).

For each cell type, we further analyzed the differentially expressed genes (DEGs) between COVID- 19 patients and healthy individuals, and we then performed GO (biological process) and KEGG enrichment analyses using these DEGs (Figure 1C-D, Figure S2B and S3B, Table S2-S7). In the case of nasal epithelial cells, many genes were upregulated in the ciliated cells of patients with mild, moderate, and severe COVID-19, and they showed enrichment in some functions and pathways, such as COVID-19, oxidative phosphorylation, protein targeting, and viral gene expression (Table S5-S6). Surprisingly, we found that squamous cells showed the highest number of downregulated genes (200 genes) during mild/moderate COVID-19, and goblet cells displayed the highest number of upregulated genes (1,033 genes) during severe COVID-19 (Figure 1C). The 200 deregulated genes were associated with regulating translation, the cellular amide metabolic process, and RNA splicing, while the 1,033 upregulated genes were associated with the ATP metabolic process, interleukin-1- mediated signaling pathway, Wnt signaling pathway, planar cell polarity pathway, response to decreased oxygen levels, and the viral life cycle (Figure 1C and Figure S2B). Likewise, we found that compared to the number of upregulated genes, a larger number of genes tended to be downregulated in most cell types from PBMCs, and the highest differences between healthy and severe COVID-19 patients were noted in CD14 monocyte cells (Figure 1C). During severe COVID- 19, upregulated genes in both CD14 and CD16 monocyte cells were associated with type I interferon signaling, response to the virus, positive regulation of cytokine production, and toll-like receptor (TLR) signaling pathways (Figure 1D). TLRs have been reported to play an important role in responses to certain infections, and their changes may lead to cytokine storms (14,40). We also found that I-kappaB kinase/NF-κB signaling was enriched. A recent study suggested that NF-κB might be associated with a poor pro-inflammatory cytokine production mechanism in the monocytes of patients severe COVID-19 (32). Other findings based on the GO and KEGG analyses among cell types are given in Table S7. These results may provide an important reference for understanding the mechanisms of cytokine storms in different cell types.

By integrating respiratory epithelial and peripheral immune cells, the SCENIC analysis identified similar and dissimilar TFs between the conditions. Furthermore, different combinations of these TFs were common or unique to certain cell types under different conditions. These distinct cell type- or condition-specific TFs profoundly contribute to transcriptional regulation among cell types during disease progression. The studies on DEGs indicated different response mechanisms to SARS-CoV-2 occur at different infection sites (based on nasal and peripheral blood data). Specifically, DEGs of certain cell types were enriched in pathways such as the regulation of cytokine production, response to decreased oxygen levels, and viral response.

### 3.3 The construction of a gene regulatory network from nasopharyngeal swabs and PBMCs evidences immune responses to SARS-CoV-2 infection in different cell types

With our identified cell type- and condition-specific regulons, further studies on TFs and their direct targets were conducted with the aim of exploring the detailed mechanisms of immune responses at different infection sites. Our findings suggested that goblet and squamous cells among epithelial cells, as well as CD14 and CD16 monocyte cells among immune cells, exhibited considerable differences in not only DEGs but also regulons. We constructed and visualized GRNs (i.e., regulons) per cell type using the Cytoscape tool (36). The target genes of each TF were further filtered using the corresponding DEGs in relation to cell types and conditions. We hoped to construct GRNs for all cell types and conditions; however, some cases failed because there were no remaining target genes of certain TFs after DEG filtering (i.e., the targets were not differentially expressed and thus the regulons were removed), or all of the regulons of these cell types or conditions showed very low activation. As a result, the GRNs of goblet, squamous, CD14, and CD16 monocyte cells, were constructed (Figure 2). In goblet cells, STAT2 and KLF5 were highly activated and upregulated in many genes with mild/moderate COVID-19, such as ISGs (PARP14 and IFI44L), whereas ELF3, SBP1, NR2F6, and SPDEF were preferentially activated in severe COVID-19 to regulate the expression of their targets (Figure 2A). We found that related TFs were expressed, and regulons showed high AUC scores in the corresponding cell types (Figure 2B and Figure S4). Furthermore, their target genes, including cytokines, interferon-stimulated genes (ISGs), and S100/Calbindin genes, were significantly upregulated in severe COVID-19 patients (Figure 2C). For squamous cells, there were no activated regulons in SPEDEF and ELF3 in mild/moderate COVID-19, but they were activated in severe COVID-19 (Figure 2A). In summary, SPEDEF and ELF3 were shared between goblet and squamous cells in severe COVID-19, and S100A9 was co-upregulated. The regulators of S100A8 were not identified, although they were significantly upregulated in the two cell types.

**Figure 2.**
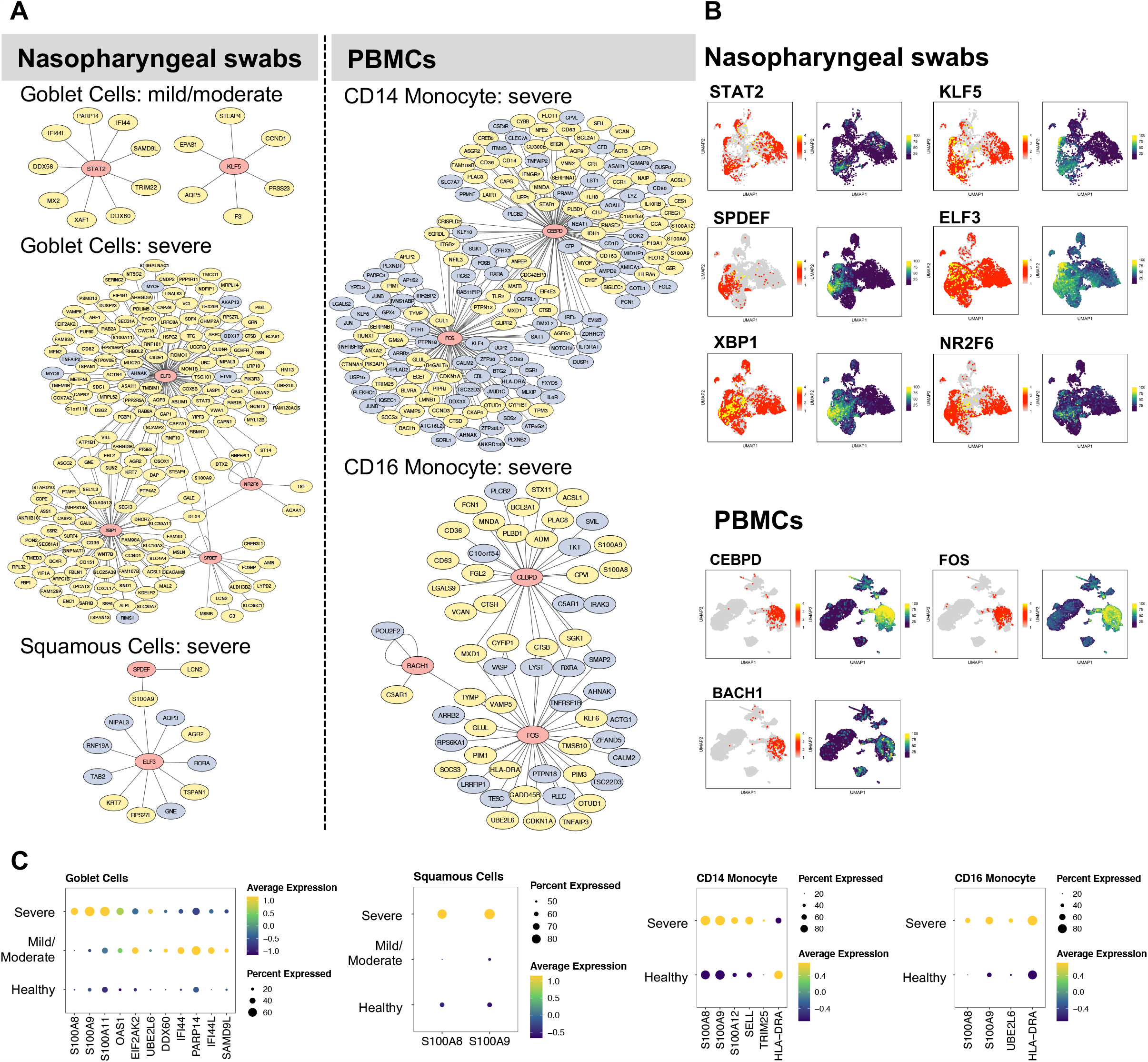
Gene regulatory networks of specific cell types. (A) The gene regulatory networks (GRNs) of specific cell types from COVID-19 patients were constructed using Cytoscape software. In the network, the red, yellow, and light blue colors represent transcription factors, upregulated, and downregulated genes, respectively. (B) UMAP plot visualization of TF expression (left) and AUC score (right) of each regulon for nasopharyngeal swabs and PBMCs data. (C) The dot plots represent target gene expressions of detected TFs in (B). The target genes include specific cytokines, interferon-stimulated genes (ISGs), and inflammatory genes (S100A8, S100A9).

However, MAFF and GRHL1 downregulated S100A8 and S100A9, respectively, in healthy squamous cells (Figure S5). In addition, ELF3 upregulated the expression of S100A11 in goblet cells (Figure 2). Similarly, for CD14 and CD16 monocyte cells in severe COVID-19, CEBPD and FOS were particularly activated in the two cell types, and CEBPD upregulated S100A8 and S100A9 (Figure 2). We also identified the substantial expression of SELL (an ISGs) in most CD14 monocyte cells in severe COVID-19. BACH1 was uniquely regulated in CD16 monocytes. Unexpectedly, HLA-DRA, a major histocompatibility complex II (MHC-II) molecule, was considerably downregulated in CD14 monocyte cells, but upregulated in CD16 monocyte cells. MHC I molecules are considered to contribute to the SARS-CoV infection response (40). A recent study demonstrated that epithelial cells with SARS-CoV-2 RNA+ express only MHC-I and poorly express MHC-II family genes (21). However, a previous study discovered that the MHC class II transactivators, CIITA and CD74, can defend against many viruses, such as SARS-like coronaviruses; therefore, upregulation of MHC-II family genes may block the entry of viruses (41).

We also constructed GRNs in other respiratory epithelial and peripheral immune cells, such as ciliated, B, T, and NK cells (Figure S5 and S6). Notably, although certain TFs were shared among cell types and conditions, their target genes and regulations differed considerably. For example, RFX2 and RFX3 were activated in ciliated cells under all three conditions, but the majority of targets were downregulated in mild/moderate COVID-19 patients compared to healthy cells (Figure S5).

Furthermore, RFX3 upregulated most of its target genes in patients with severe COVID-19. For the NK cells, many ISGs were upregulated by STAT1 in severe COVID-19, such as EIF2AK2, PARP14, ISG15, PSMB9, MX1, SP110, DDX60, SAMD9L, ADAR, IFI44L, IFIT3, EPSTI1, SAMD9 (Figure S6). In patients with severe disease, we also found that TCF4 was activated in B cells, whereas RUNX3, IKZF1, and EOMES were activated in CD8 cells (Figure S6).

By integrating different cell types from nasal or peripheral blood during progression to severe COVID-19, our findings demonstrated the existence of diverse GRNs. Intriguingly, we found that S100A8 and S100A9 were considerably upregulated by different TFs in a wide range of respiratory epithelial and peripheral immune cells in patients with severe COVID-19 (Figure S4), which suggests that their upregulation tends to be independent of certain cell types and virus-infection sites but that different regulators can be used among cell types. Specifically, the systemic upregulation of S100A8 and S100A9 mainly occurred in goblet, squamous, B, CD14/CD16 monocytes, granulocytes, PB, and DC. However, their regulators can differ in terms of cell type at different infection sites. S100A8 and S100A9 have been reported to be markers of severe COVID-19 (18) and contribute to the recruitment of immune cells and cytokine storms in megakaryocytes and monocytes (21,33,42).

### 3.4 Robust DEGs were identified in T cells

To identify genes robustly expressed during the immune response against SARS-CoV-2 infection, we further observed the overlap of DEGs in T cells from nasal and PBMC scRNA-seq data. Compared to one type of T cell in the nasal data, three types of T cells were annotated in PBMC data: CD4, CD8, and gd T cells. We extracted corresponding DEGs when comparing healthy individules and severe patients to identify overlapping DEGs by comparing one T cell type from nasal data to three T cell types from PBMC data, and then visualized their expressions in the two datasets (Figure 3). The majority of overlapping genes were robustly downregulated in patients with severe disease, but only one overlapping gene, prothymosin alpha (PTMA), was consistently upregulated. Interestingly, PTMA, the proprotein of thymosin alpha-1 (Tα1), has been reported to show increased expression in CD8 T memory cells in severe disease and slightly reduced activation of T cells in vitro (43), and the authors indicated that lymphopenia in COVID-19 patients could be relieved by Tα1 treatment.

**Figure 3.**
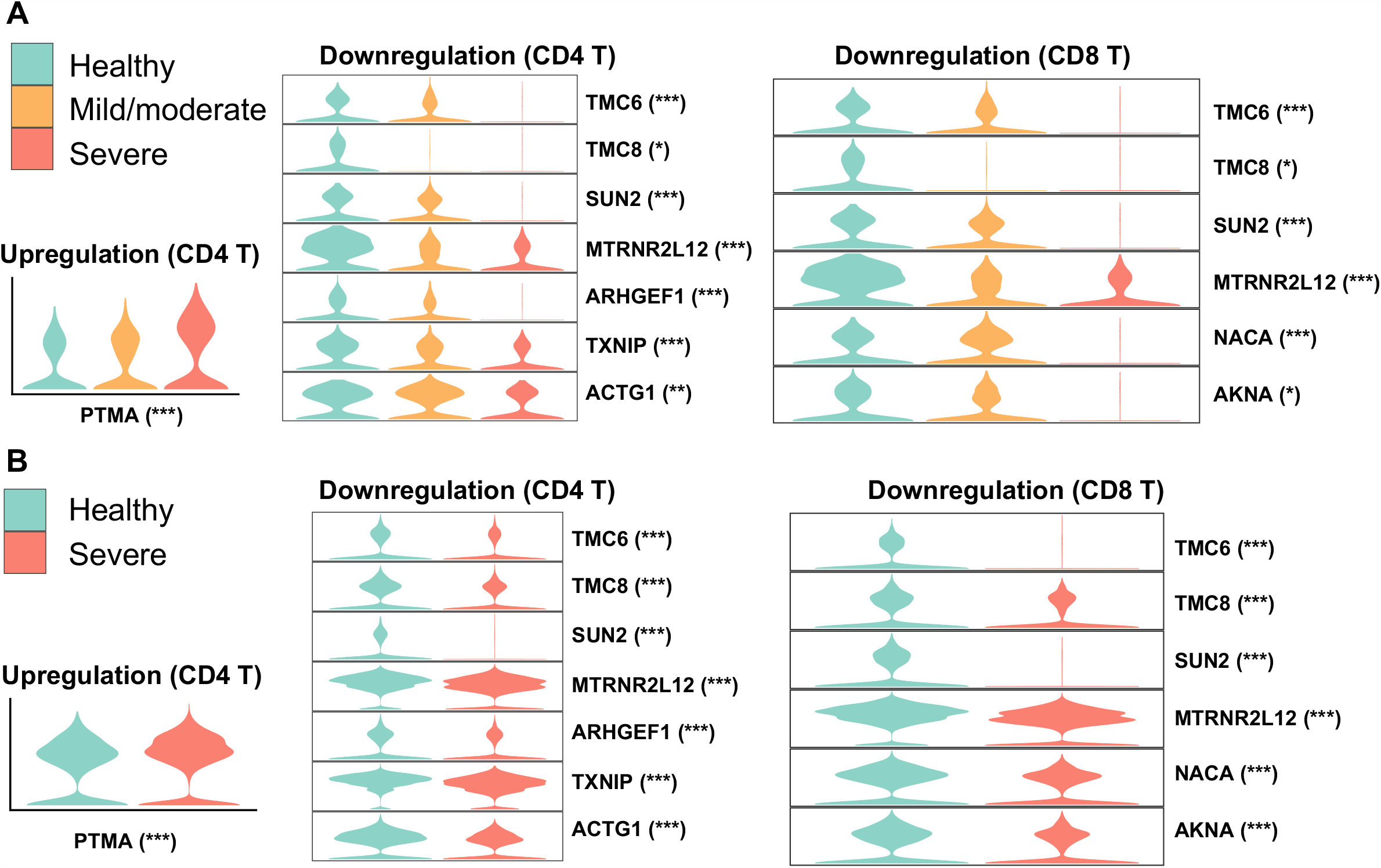
Overlap of DEGs in nasal and PBMC T cells when comparing healthy and severe COVID-19 cases. Violin plots represent expressions of overlapping DEGs in (A) nasopharyngeal swabs and (B) PBMC data. The asterisk following each gene indicates a significant difference between healthy and severe COVID-19 cases. Significance: * P < 0.05, ** P < 0.01, *** P < 0.001.

Among the overlapping downregulated genes, genetic defects in TCM6/8 may lead to lower intrinsic immunity to human β-papillomaviruses (β-HPVs) in epidermodysplasia verruciformis patients (44). SUN2 (Sad1 and UNC84 domain containing 2) is associated with mitosis, it maintains a repressive chromatin state, and inhibits HIV-1 infection via association with Lamin A/C (45,46). Previous studies have also shown that quercetin and resveratrol, inhibitors of thioredoxin-interacting protein (TXNIP), could be considered as potential therapies for COVID-19 (47,48). However, we found that TXNIP showed robust downregulation in severe patients, which suggests that its inhibitors may provide a reduced performance in severe patient treatment to a certain degree.

### 3.5 Alterations in enriched pathways and cell-cell communications between mild/moderate and severe COVID-19 were identified

As nasal epithelial cells mute the antiviral response in severe COVID-19 compared to mild/moderate patients, and this early failure may underlie and predict severe COVID-19 (21), we analyzed and compared epithelial cells between mild/moderate and severe COVID-19 patients. There was a greater upregulation in squamous and goblet cells in patients with severe disease than in those with mild/moderate (Figure 4A). Upregulated genes in squamous cells were associated with pathways such as viral entry into the host cell, epidermis development, protein localization, and apoptotic signaling, while genes in goblet cells were related to protein targeting, response to hypoxia, and viral gene expression (Figure 4A). Although only a few downregulated genes were identified in squamous (19 genes) and goblet (50 genes) cells, they were still enriched in certain pathways. For example, CD74, TNFAIP3, and S100A4 in squamous cells were associated with I-kappaB kinase/NF-kappaB signaling and interleukin-6 production, while MX1, IFIT1, SP100, and XAF1 in goblet cells were associated with the type I interferon signaling pathway (Figure S7).

**Figure 4.**
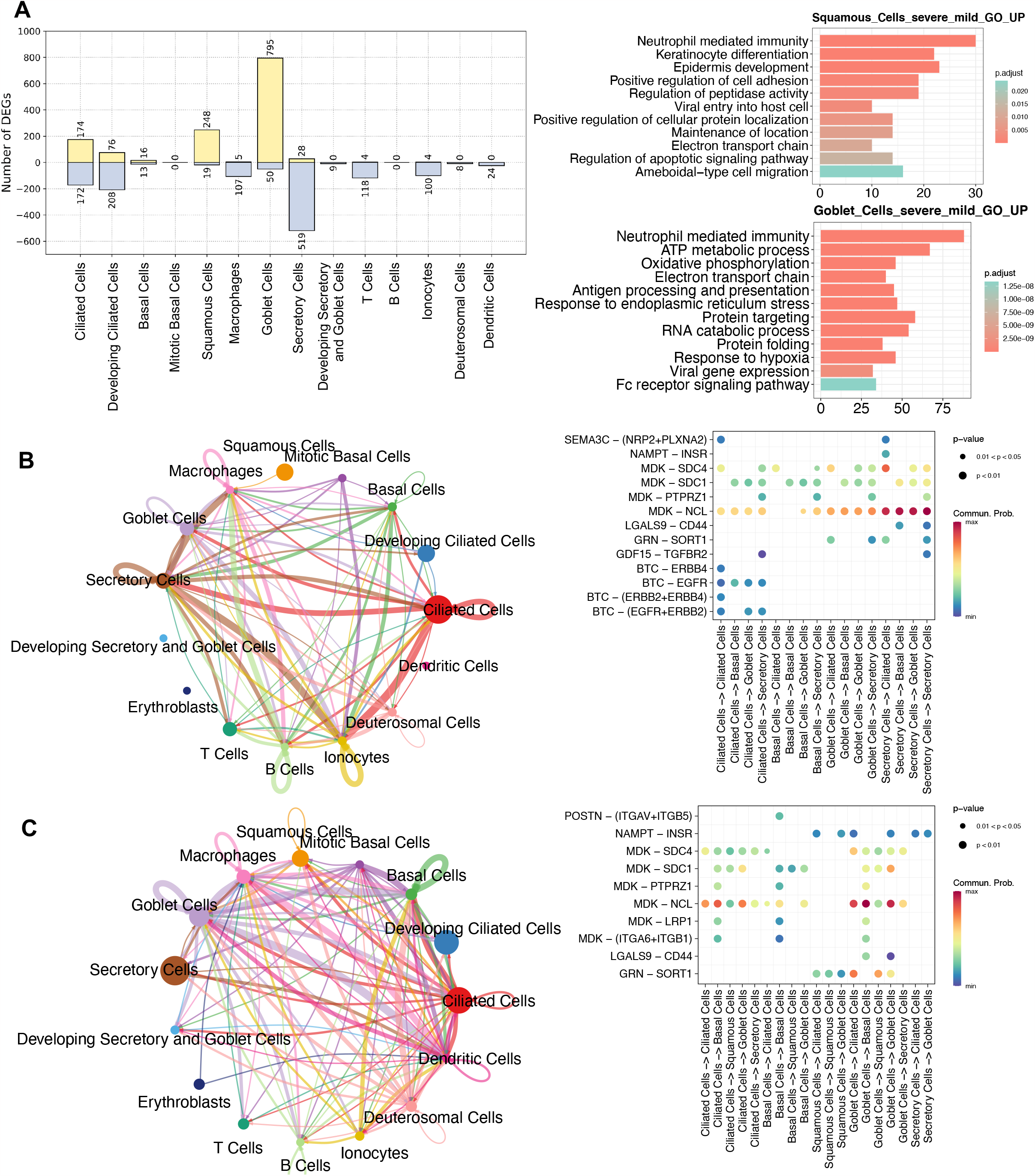
Comparisons between mild/moderate and severe COVID-19 from nasopharyngeal swabs. (A) Number of identified DEGs when comparing severe and mild/moderate COVID-19. Yellow and light- blue colors represent upregulated and downregulated genes, respectively. The right panel indicate the results of GO enrichment analysis using DEGs based on biological process. Cell-cell communication (CCC) in (B) mild/moderate and (C) severe COVID-19. The left-hand networks of (B) and (C) represent cell interaction numbers among cell types. Cell types are represented by circles with different colors. Circle size indicates the number of cells of a cell type, while the edge width corresponds to the numbers of ligand-receptor (L–R) interactions. The Bubble plots (right-hand side of B and C) show all the significant L–R pairs associated with signaling pathways from a given cell type to another one. The dark blue to red colors relate to the communication probability (from minimum to maximum).

To investigate the ligand-receptor (L–R) interactions among epithelial cells, we performed a cell-cell communication (CCC) analysis using CellChat (38). Compared to patients with mild/moderate COVID-19, our CCC analysis suggested that epidermal growth factor receptor (EGFR) signaling from ciliated cells to other cell types was lost in patients with severe COVID-19 (Figure 4B-C).

EGFR (also known as ErbB1) belongs to a family of receptor tyrosine kinases (ErbB), and ErbB contains four receptors: ErbB1, ErbB2, ErbB3, and ErbB4 (49). In this study, three of the receptors (not ErbB3) were identified in mild/moderate patients. The ligand of these receptors is betacellulin (BTC), and a previous study indicated that it might be useful in preventing an excessive fibrotic response to viral infections (such as SARS-CoV) by inhibiting EGFR signaling (50). Similarly, we observed an absence of EGFR signaling in healthy cells (Figure S8A), and we therefore consider that EGFR inhibitors could be used as a potential treatment for mild/moderate COVID-19. In contrast, L–R interactions associated with the midkine (MDK) signaling pathway were observed in ciliated, secretory, and other cells in healthy, mild/moderate, and severe COVID-19 patients (Figure 4B-C and Figure S8A).

### 3.6 Whole blood bulk transcriptomic data analysis showed genes correlated with the sequential organ failure assessment score

To test whether our identified genes could contribute to immune responses during the progression of COVID-19 severity, we downloaded public whole blood bulk transcriptomic data of COVID-19 patients for validation (39) and analyzed alterations in gene expression levels in terms of the sequential organ failure assessment (SOFA) score. The transcripts per million (TPM) method was used to normalize the transcript counts. We then calculated and ranked the Pearson correlations between every single gene TPM value and the SOFA scores in COVID-19 patients (Table S9). We found that 26 genes were highly positively correlated with SOFA scores (Pearson’s r > 0.50) (Figure 5A), indicating that gene expression levels increased with the severity of clinical organ failure in critically ill patients. Intriguingly, among the top 26 correlated genes, two were from the S100 gene family: S100A8 and S100A12. According to our scRNA-seq data findings in this study, these two genes were differentially expressed in patients with severe COVID-19 (Figure 2C). We further calculated the gene average TPM value for each SOFA score, including several S100 and otherN genes, as shown in Figure 5A (Figure 5B). The S100A8/A12 expression increased with an increasing SOFA severity score. The expression of S100A9 showed a certain correlation with the SOFA score (*r = 0*.*22*), but S100A11 was negatively correlated. Notably, we found that matrix metalloproteinase family members (MMPs), MMP8 and MMP27, were highly correlated. It has been reported that MMPs contribute to the COVID-19 severity (51–53). Taken together, our findings indicate the critical roles of S100 family members and other genes (e.g., MMPs) in the progression of COVID- 19.

**Figure 5.**
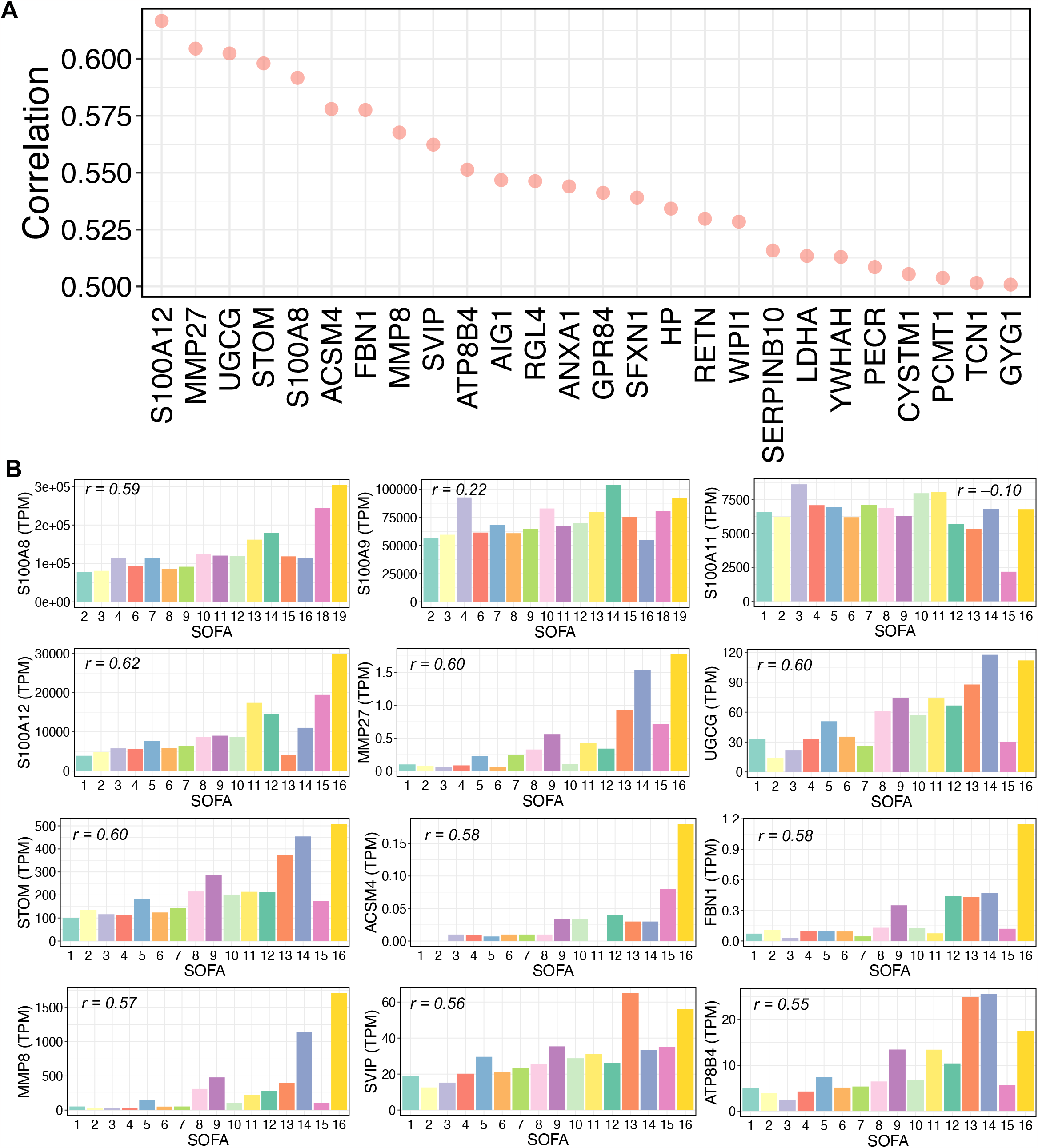
Relationships between gene expressions and SOFA scores in COVID-19 samples. (A) Pearson correlations between gene expression and sequential organ failure assessment (SOFA) score. Top 26 genes with over 0.5 correlations are shown. Transcripts per million (TPM) were used for RNA-seq normalization. (B) Average TPM values of selected genes for each SOFA score. The selected genes include some S100 family members and some genes from the top 26 gene list in (A). For each bar plot, the Pearson’s “r” between the TPM values and SOFA scores of all related samples is shown.

## 4 Discussion

SARS-CoV-2 infection causes immune response alterations during the progression of COVID-19 severity, such as the enhanced expression of pro-inflammatory cytokines (i.e., cytokine storm) and considerably different expressions of genes associated with ISGs, MHC I, and II families (11–13,21,24,41,54). However, the GRN changes in terms of a wide range of cell types and COVID-19 severity (healthy, mild/moderate, and severe) require further description. In this study, therefore, we conducted a SCENIC analysis to capture similar and dissimilar regulons, and we then finely constructed GRN landscape for both healthy individuals and COVID-19 patients across a wide range of cell types by integrating scRNA-seq data relating to the site of infection (respiratory epithelial cells) and peripheral blood (peripheral immune system). Further analyses of the target genes of regulators and cell-cell communication revealed detailed intracellular and extracellular immune responses. Lastly, the comparison of findings using different data sources and the validation of certain key findings using bulk RNA-seq data provided greater robustness to our results.

We identified cell type- and condition-specific activated regulons (including TFs and their targets) in a wide range of cell types, and we demonstrated that some cells, such as goblet, squamous, and monocyte cells, display a strong response against SARS-CoV-2 infection. For example, compared to activated STAT2 and KLF5 in mild/moderate goblet cells, SPDEF, ELF3, XBP1, and NR2F6 were found to be activated in patients with severe COVID-19. Similarly, SPDEF and ELF3 were found to be activated in squamous cells, and CEBPD and FOS were shared between CD14 and CD14 monocytes in severe COVID-19. We then constructed and compared GRNs. Most importantly, by integrating nasopharyngeal swab and PBMCs data, we discovered the regulation of S100A8 and S100A9 expression, which could help to clarify maladaptive responses to SARS-CoV-2 infection in a large range of cell types. Specifically, we found that SPEDEF and ELF3 co-upregulated S100A9 in goblet and squamous cells in severe patients, whereas CEBPD upregulated S100A8 and S100A9 in monocyte cells. In contrast, S100A8 and S100A9 were downregulated by MAFF and GRHL1 in healthy squamous cells. We also observed that during severe COVID-19, S100A8 and S100A9 were considerably upregulated in multiple other cell types, such as B cells, granulocytes, PB, and DC. The expression level of S100A8 was further found to have a high positive correlation with the SOFA score (Figure 5). A previous study reported the same regulation in T, B, NK, and DC cells (33).

However, compared to healthy individuals, we did not observe upregulation of S100A8 and S100A9 in CD4, CD8, and gd T cells in severe patients. In addition, we identified regulators of many cytokines, ISGs, or MHC II family genes in severe patients, such as gene regulations of goblet (CLCL17, EIF2AK2, UBE2L6), monocyte (HLA-DRA, SELL, TRIM25, UBE2L6), CD8 T (SAMD9), NK (EIF2AK2, SAMD9, DDX60, PARP14, MX1, PARP9, ADAR, TXNIP, ISG15, IFIT3, EPSTI1, SP110, SAMD9L, IFI44L, TAP1), granulocyte (RSAD2, MX1, IFI44, EPSTI1, EIF2AK2, IFIT3, IFI44L, ISG15), and DC cells (MX1, RSAD2, IFI44L, IFI44, ISG15, IFIT3) (Figure 2, Figure S5, and S6). Notably, although ciliated cells play critical roles in viral entry, we did not detect distinct regulators in infected patients. Regulators RFX2 and RFX3 were shared between healthy individuals and COVID-19 patients, but the two regulators showed different target genes or target expression levels in the two sample types (Figure S5). Further CCC analysis demonstrated that compared to severe patients, EGFR signaling has an important role when ciliated cells interact with other cells in mild/moderate patients.

We also used PBMC data to investigate cell-cell interactions among immune cells. The chemokine (C-C motif) signaling pathway was found to be unique to severe patients compared to healthy individuals (Figure S8B-C). This pathway mainly contributes to pathways from CD8 T and NK cells to CD14 monocytes via the CCL5 and CCR1 ligand-receptor pairs. A recent study demonstrated that CCL5 contributes to the recruitment of inflammatory cells (mainly T cells and macrophages), and blockading CCR5 signaling using leronlimab (a monoclonal antibody) was conducted to treat COVID-19-associated cytokine storms (55).

In summary, this study integrated healthy individuals and COVID-19 patient data from two independent sources (nasopharyngeal swabs and PBMCs scRNA-seq data), compared the findings of each dataset, and validated certain key findings using bulk RNA-seq data. By separately analyzing the data from each study, we revealed the GRN landscape and identified similar and dissimilar regulons and pathways under different conditions (i.e., healthy individuals and COVID-19 patients). Regulators of certain key genes (e.g., S100A8/A9) were found to differ among cell types and with disease severity. However, when comparing and combining the two independent studies, we found that virus-infected individuals had certain common and/or unique features at different infection sites. For example, (i) certain activated regulators (such as XBP1, FOS, STAT1, and STAT2) were shared in both respiratory epithelial and peripheral blood; (ii) certain genes (e.g., PTMA) were differentially expressed in T cells from independent studies; (iii) although S100A8/A9 were found to be upregulated in both respiratory epithelial and peripheral blood, their relative regulators differed (e.g., SPEDEF and ELF3 were found in goblet and squamous cells and CEBPD was seen in monocyte cells, which showed that regulators of a gene were specific to the infection site, cell type, and condition. Collectively, the results using our approach provide clues to comprehensively understanding the diverse disease mechanisms of SARS-CoV-2 infection. These findings can be used as a rich resource for predicting, preventing, and treating COVID-19 in a wide range of cell types, which may help control severe symptoms.

This study has several limitations. The numbers of epithelial cell types were not equal: some were obtained in large quantities and others in small quantities. It is acknowledged that a small number of cells may influence transcriptional landscapes. In the case of PBMC data, we lacked those associated with mild and moderate COVID-19. Technical issues, such as those encountered with scRNA-seq techniques, have led to certain limitations that may be difficult to overcome. Nonetheless, by integrating additional information and data from independent sources where available, some of these limitations may be resolved. Therefore, further studies using large-scale data and finer-grained descriptions of COVID-19 severity categories would help to fully understand GRNs, their gradual shift, and dynamics in terms of COVID-19 severity progression. Future characterizing the specific role of some genes (e.g., S100 family members) in the immune response to SARS-CoV-2 infection, particularly in relation to disease severity, is needed in vitro and vivo experiments.

## 5 Data availability statement

The datasets presented in this study can be found in the online repositories. The names of the repository/repositories and accession numbers (s) can be found in the article/supplementary materials.

## 6 Author contributions

LZ conceived and designed the study. LZ contributed to data collection and analysis. LZ, HN, and KK interpreted the results. LZ wrote the original manuscript. HN and KK reviewed and edited the manuscript. All the authors contributed to the article and approved the submitted version.

## Data Availability

All data produced in the present work are contained in the manuscript

## Supplementary material

**Supplementary Figure S1.**
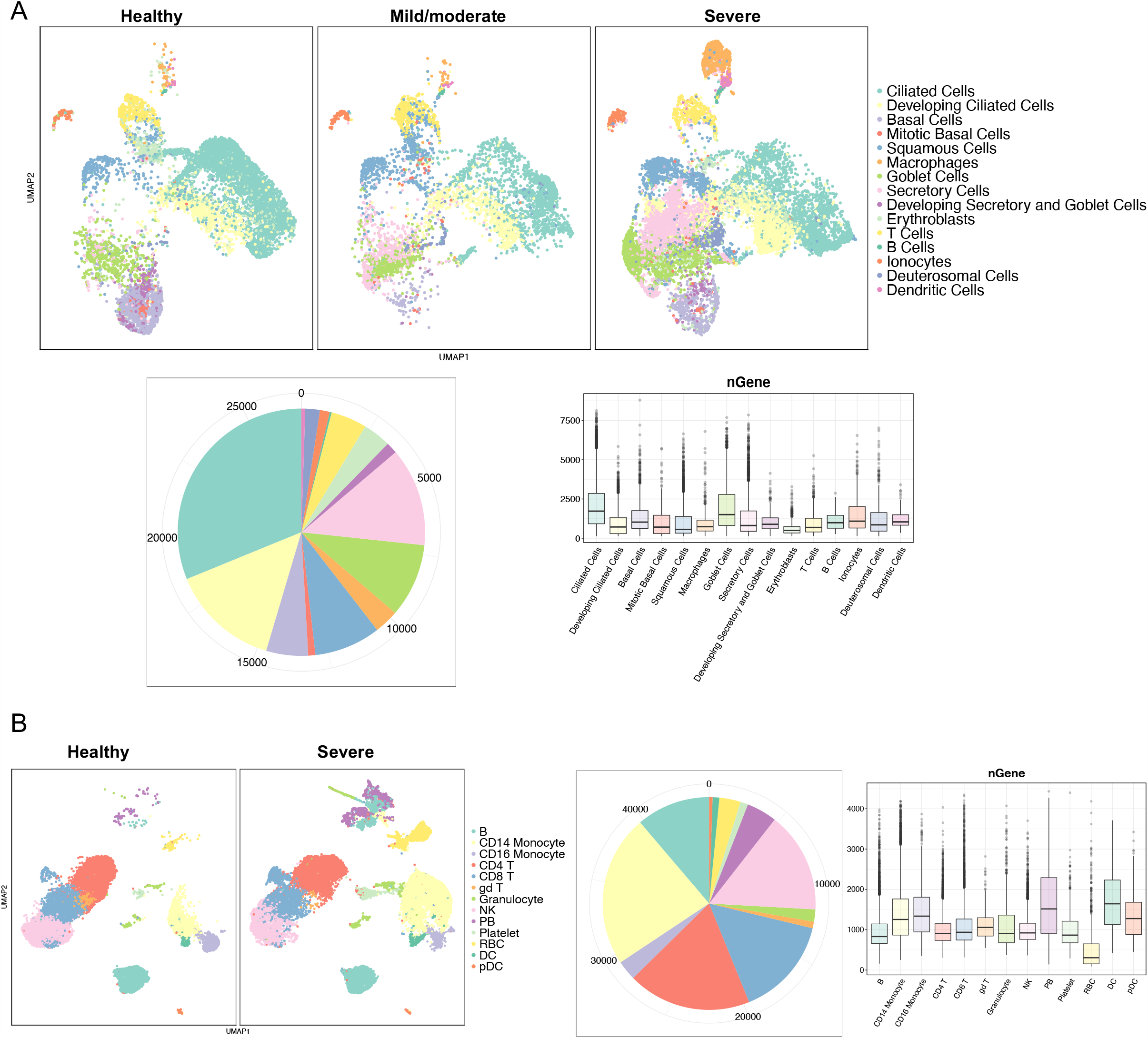
Characterization of (A) nasopharyngeal swabs and (B) PBMCs. UMAP plots showing cell types associated with condition (healthy, mild/moderate, and severe). The pie plots indicate the total number of cells used in the study, and the bar plots show the number of genes in each cell type. The lower and upper hinges of the box represent the first and third quartiles (25th and 75th percentiles), the median is marked within the box, and the dots represent outliers. Upper whisker = 75th percentile + 1.5* interquartile range (IQR). Lower whisker = 25th percentile – 1.5*IQR.

**Supplementary Figure S2.**
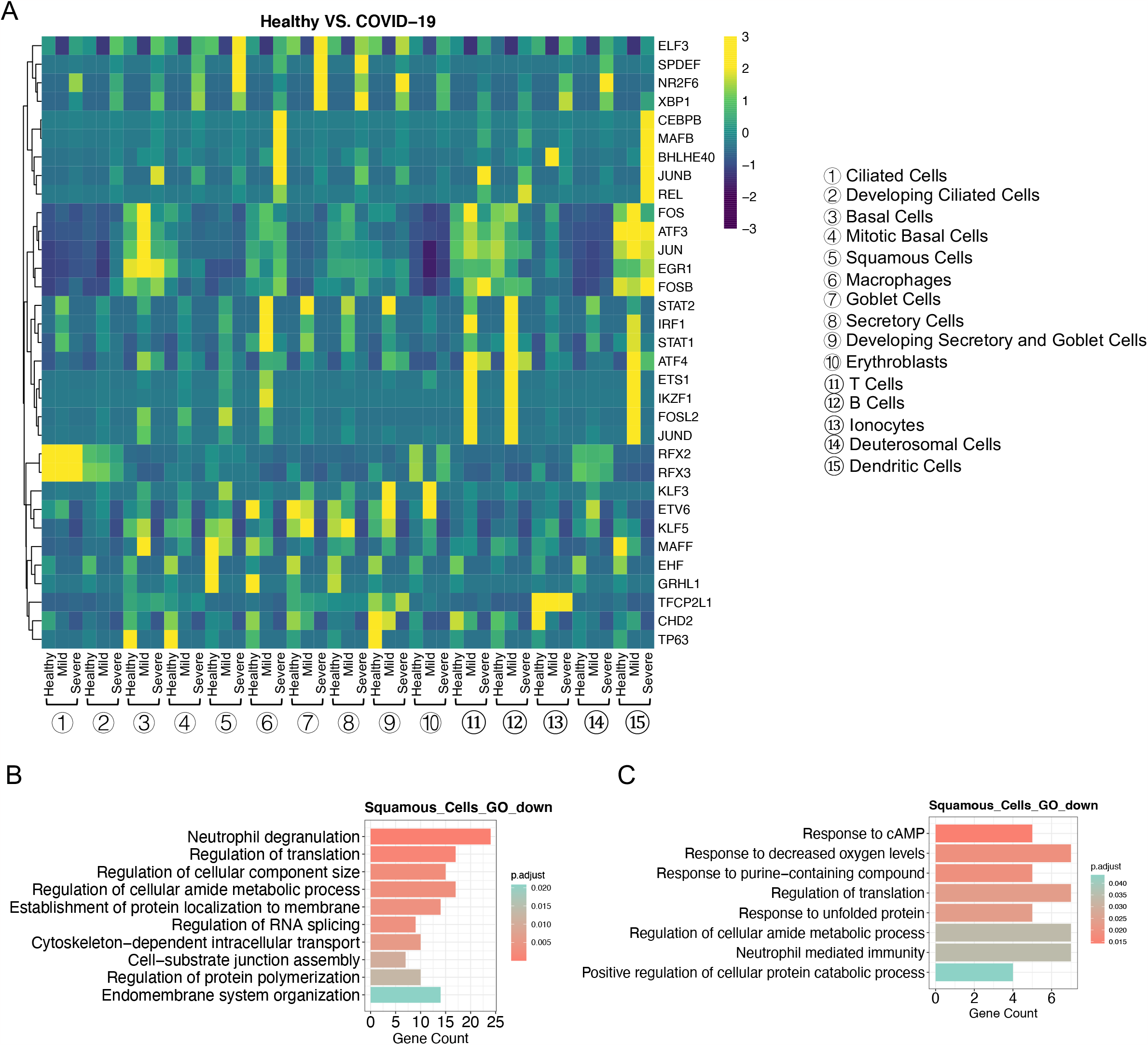
Characterization of nasopharyngeal swabs data. (A) Heatmap of area under the curve (AUC) scores of regulons estimated per cell type by SCENIC. Detected regulons are represented by their corresponding transcription factors in the right-hand columns. (B) GO (biological process) enrichment analysis using downregulated genes to compare mild/moderate to healthy cells. (C) GO enrichment analysis using downregulated genes to compare severe to healthy cells.

**Supplementary Figure S3.**
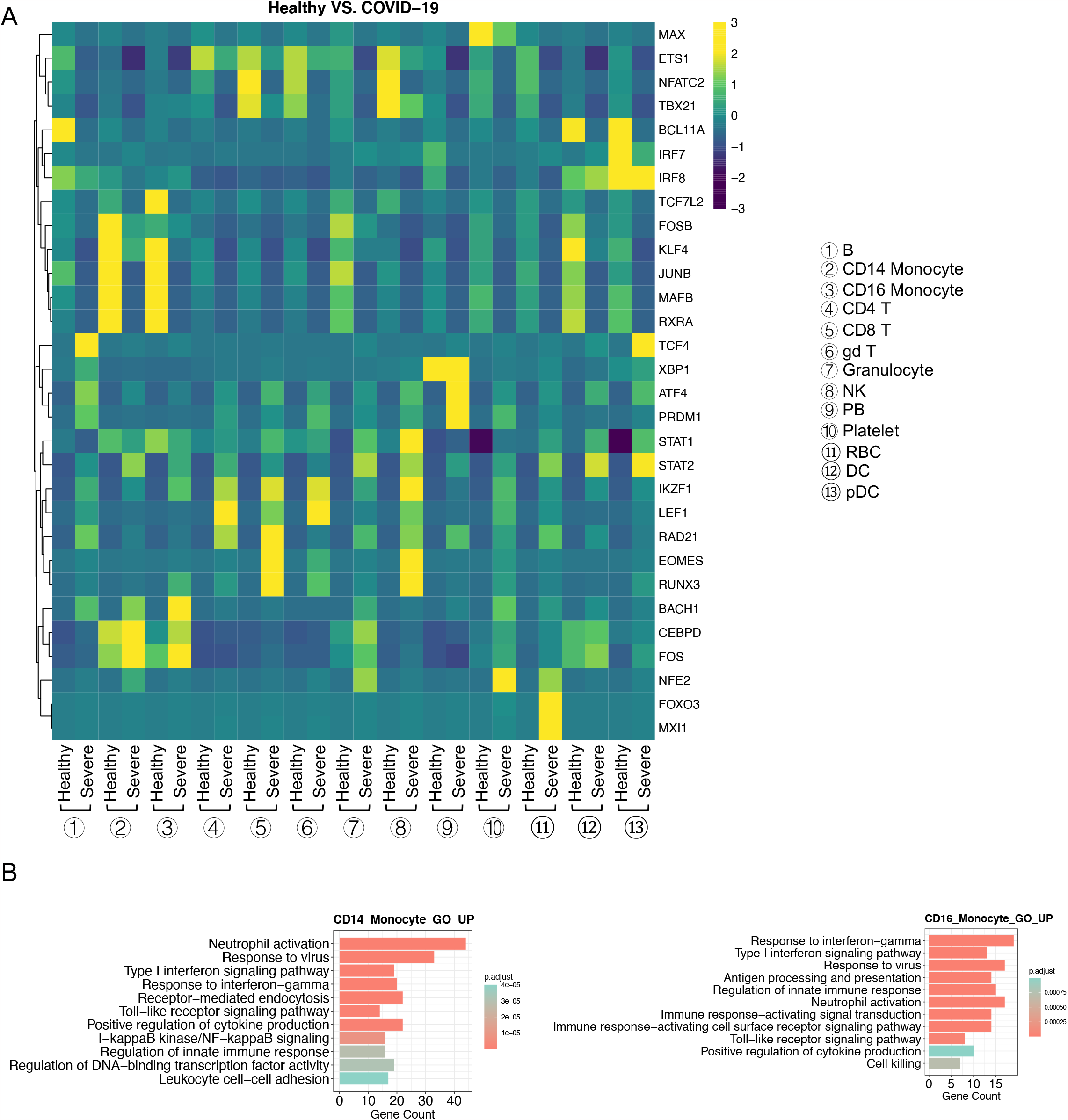
Characterization of PBMC data. (A) Heatmap of area under the curve (AUC) scores of regulons estimated per cell type by SCENIC. Detected regulons are represented by their corresponding transcription factors in the right-hand columns. (B) GO enrichment analysis using downregulated genes to compare severe to healthy cells.

**Supplementary Figure S4.**
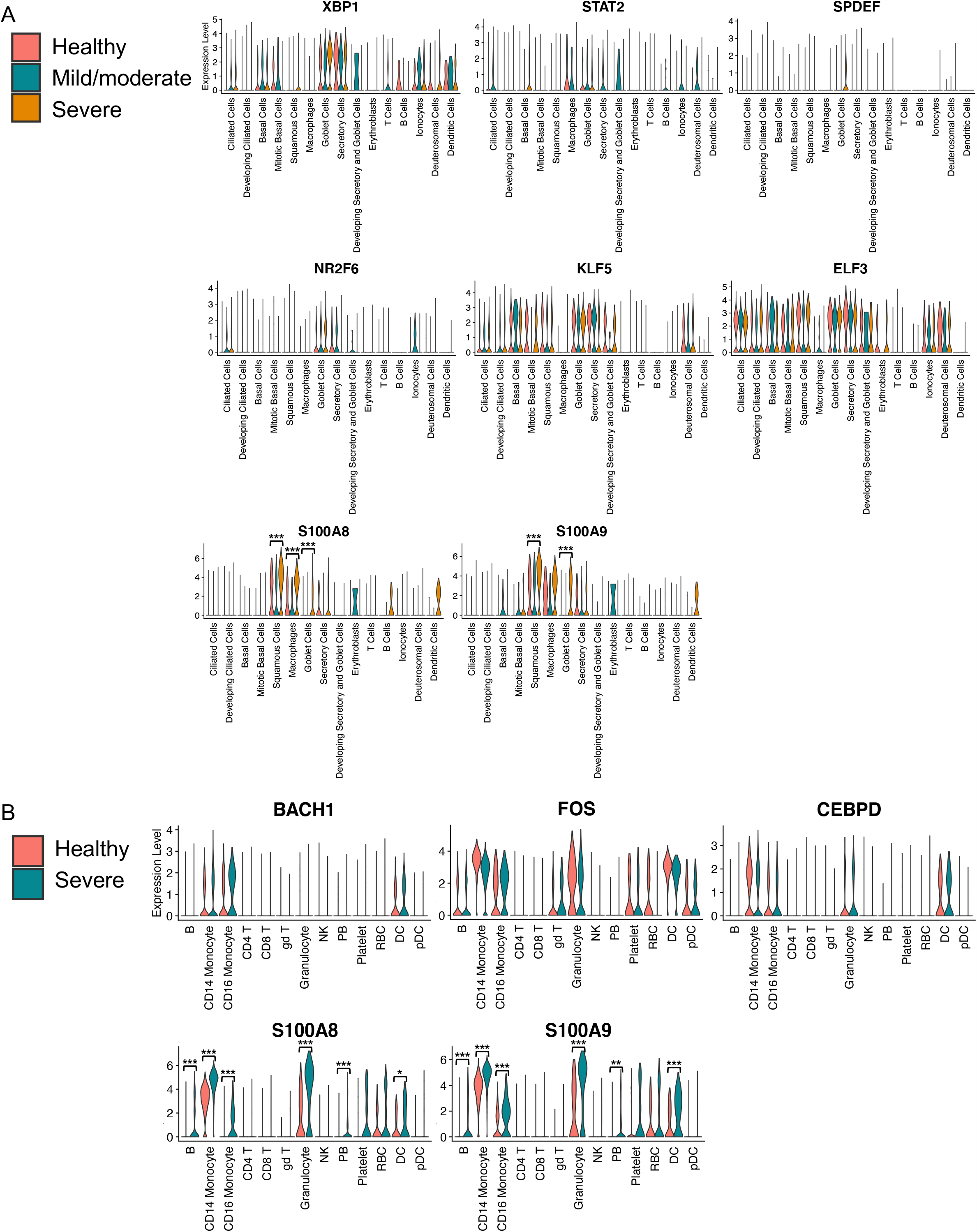
Expressions of selected genes for nasopharyngeal swabs (A) and PBMC (B) data. The violin plot shows gene expressions across cell types and conditions (healthy, mild/moderate, and severe COVID-19). Significant expression levels of S100A8/A9 when comparing healthy and severe COVID-19 patients are shown as follows: * P < 0.05, ** P < 0.01, *** P < 0.001.

**Supplementary Figure S5.**
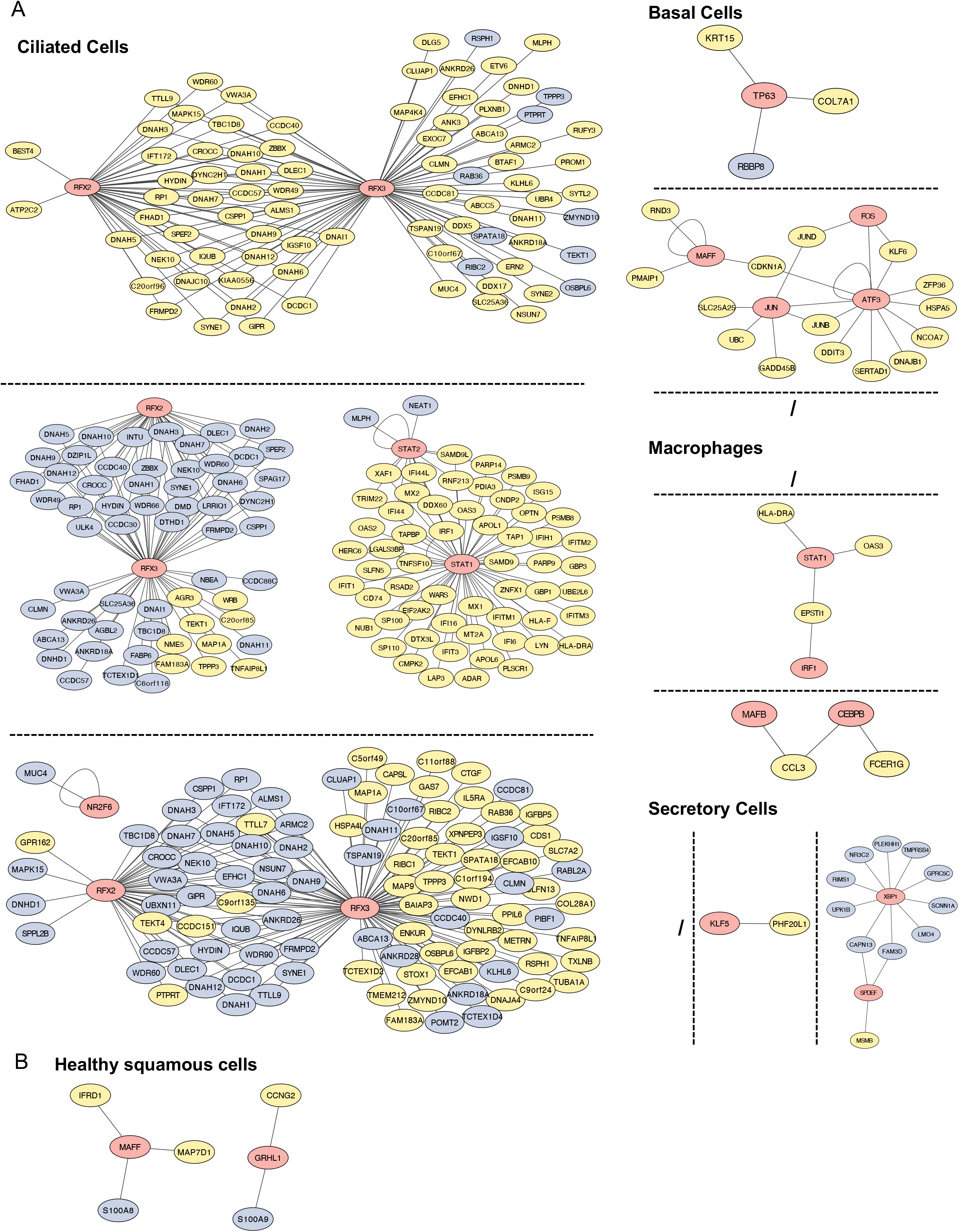
Gene regulatory networks of cell types from nasopharyngeal swabs. (A) Each panel shows the GRNs in healthy, mild/moderate, and severe COVID-19 from top to bottom, respectively. The slash symbol represents no observed regulons or DEGs. (B). GRNs of healthy squamous cells. In the network, the red, yellow, and light-blue colors represent TF, upregulated, and downregulated genes, respectively.

**Supplementary Figure S6.**
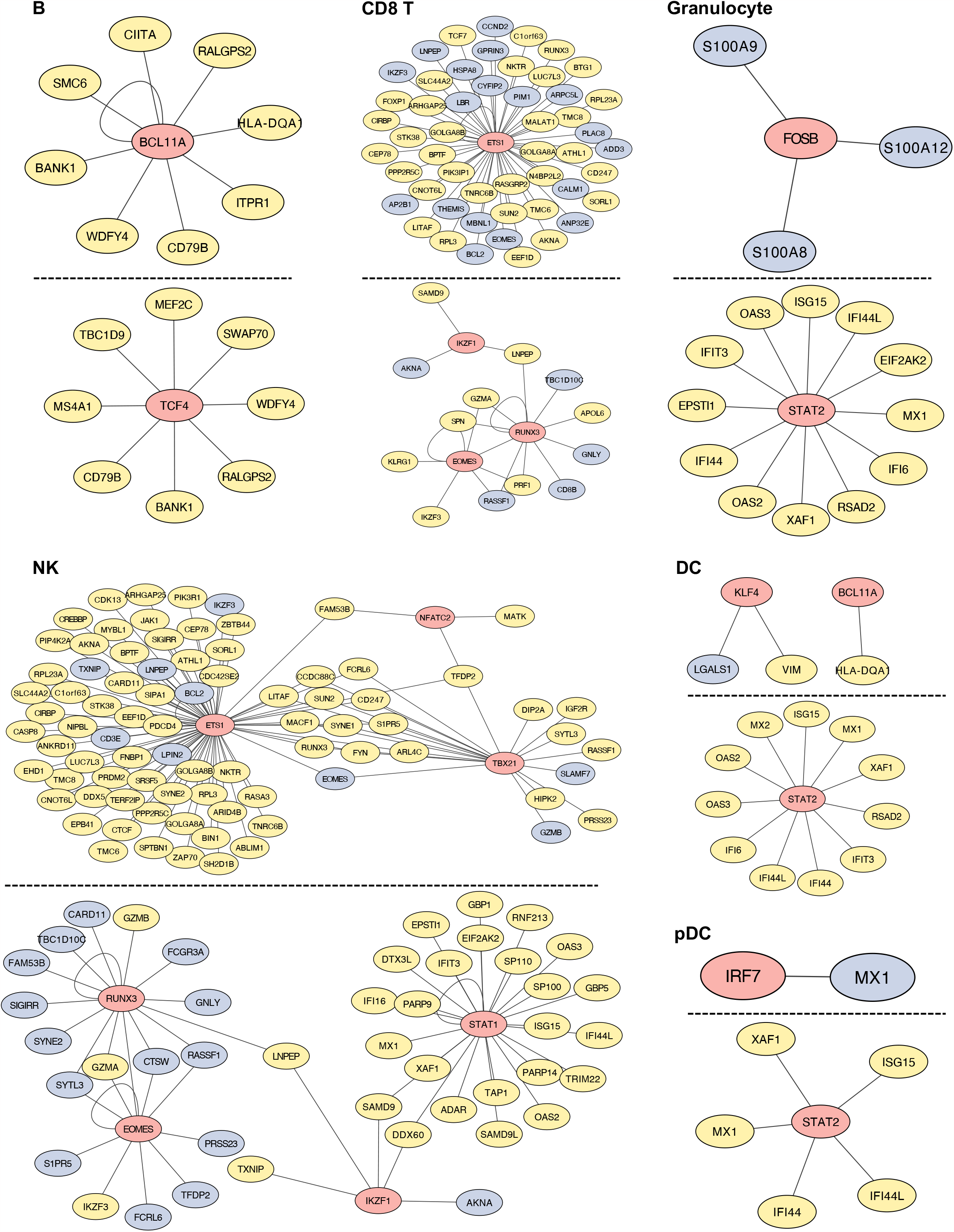
Gene regulatory networks of cell types from PBMCs. Each panel represents the GRNs in healthy and severe COVID-19 groups from top to bottom, respectively. The slashed symbol represents no observed regulons or DEGs. In the network, the red, yellow, and light-blue colors represent TF, upregulated, and downregulated genes, respectively.

**Supplementary Figure S7.**
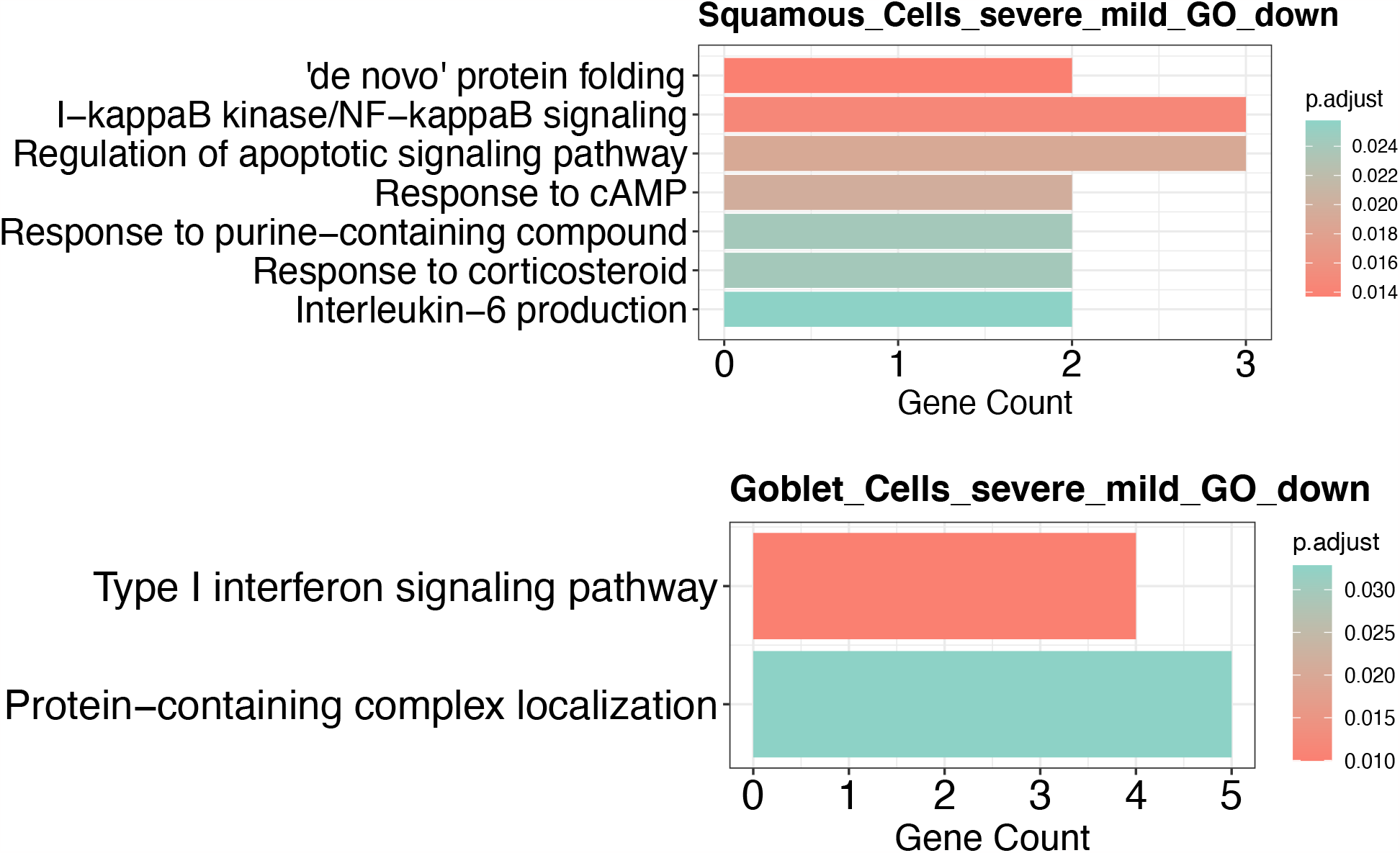
GO analysis of downregulated genes in squamous and goblet cells. The GO enrichment analysis is based on the biological processes and identified downregulated genes when comparing severe and mild/moderate COVID-19 patients.

**Supplementary Figure S8.**
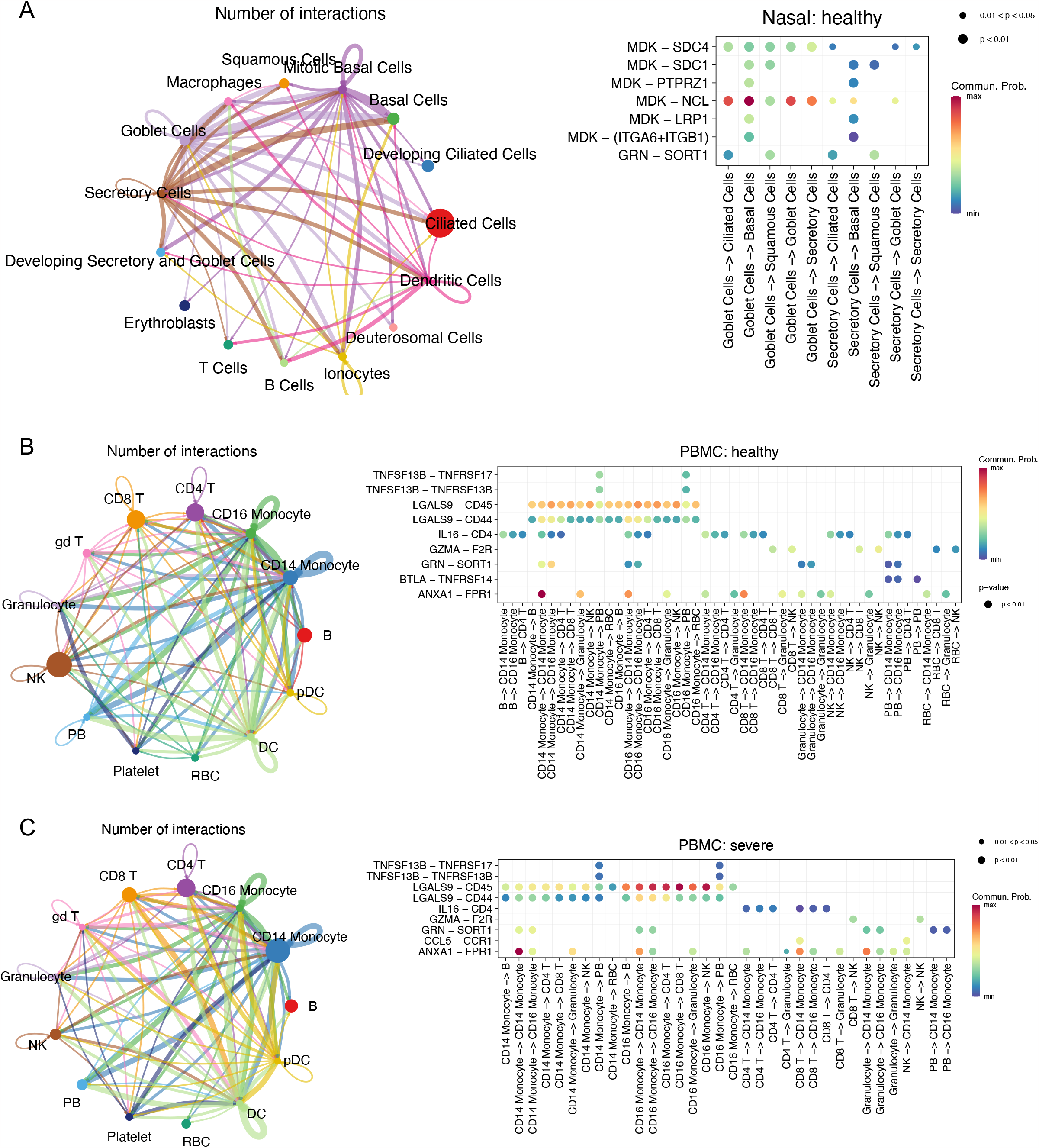
CCC analysis using nasopharyngeal swab and PBMC data. (A) CCC analysis of healthy cells from nasopharyngeal swabs. (B) and (C) represent CCC analysis of healthy and severe COVID-19 cells from PBMCs, respectively. Only significant L–R interactions associated with signaling pathways are shown in bubble plots.

## Figure legends

**Supplementary Table S1. Numbers of cell types in nasopharyngeal swabs and PBMC**

**Supplementary Table S2. DEGs of nasal epithelia comparing mild and moderate COVID-19 to healthy control across cell types**.

**Supplementary Table S3. DEGs of nasal epithelia comparing severe COVID-19 to healthy control across cell types**.

**Supplementary Table S4. DEGs of PBMCs comparing severe COVID-19 to healthy control across cell types**.

**Supplementary Table S5. GO and KEGG enrichment analyses using DEGs corresponding to Table S2**.

**Supplementary Table S6. GO and KEGG enrichment analyses using DEGs corresponding to Table S3**.

**Supplementary Table S7. GO and KEGG enrichment analyses using DEGs corresponding to Table S4**.

**Supplementary Table S8. DEGs of nasal epithelia comparing severe to mild/moderate COVID- 19 across cell types**.

**Supplementary Table S9. Ranked Pearson’s “r” between TPM values and SOFA scores using COVID-19 samples**.

## References

1. Lavezzo E, Franchin E, Ciavarella C, Cuomo-Dannenburg G, Barzon L, Del Vecchio C, Rossi L, Manganelli R, Loregian A, Navarin N. Suppression of a SARS-CoV-2 outbreak in the Italian municipality of Vo’. Nature (2020) 584:425–429.

2. Guo T, Fan Y, Chen M, Wu X, Zhang L, He T. Cardiovascular implications of fatal outcomes of patients with coronavirus disease 2019 (COVID-19). JAMA Cardiol. 2020; 5 (7): 811–8. (2020)

3. Ruan Q, Yang K, Wang W, Jiang L, Song J. Clinical predictors of mortality due to COVID-19 based on an analysis of data of 150 patients from Wuhan, China. Intensive Care Med (2020) 46:846–848.

4. Huang C, Wang Y, Li X, Ren L, Zhao J, Hu Y, Zhang L, Fan G, Xu J, Gu X. Clinical features of patients infected with 2019 novel coronavirus in Wuhan, China. Lancet (2020) 395:497–506.

5. Li G, Fan Y, Lai Y, Han T, Li Z, Zhou P, Pan P, Wang W, Hu D, Liu X. Coronavirus infections and immune responses. J Med Virol (2020) 92:424–432.

6. Cheng A, Zhang W, Xie Y, Jiang W, Arnold E, Sarafianos SG, Ding J. Expression, purification, and characterization of SARS coronavirus RNA polymerase. Virology (2005) 335:165–176.

7. Masters PS. The molecular biology of coronaviruses. Adv Virus Res (2006) 66:193–292.

8. Mason RJ. Pathogenesis of COVID-19 from a cell biology perspective. Eur Respir J (2020) 55:

9. Zhou P, Yang X-L, Wang X-G, Hu B, Zhang L, Zhang W, Si H-R, Zhu Y, Li B, Huang C-L. A pneumonia outbreak associated with a new coronavirus of probable bat origin. Nature (2020) 579:270–273.

10. Li X, Geng M, Peng Y, Meng L, Lu S. Molecular immune pathogenesis and diagnosis of COVID-19. J Pharm Anal (2020) 10:102–108.

11. Mudd PA, Crawford JC, Turner JS, Souquette A, Reynolds D, Bender D, Bosanquet JP, Anand NJ, Striker DA, Martin RS. Distinct inflammatory profiles distinguish COVID-19 from influenza with limited contributions from cytokine storm. Sci Adv (2020) 6:eabe3024.

12. Mehta P, McAuley DF, Brown M, Sanchez E, Tattersall RS, Manson JJ. COVID-19: consider cytokine storm syndromes and immunosuppression. Lancet (2020) 395:1033–1034.

13. Bhaskar S, Sinha A, Banach M, Mittoo S, Weissert R, Kass JS, Rajagopal S, Pai AR, Kutty S. Cytokine storm in COVID-19—immunopathological mechanisms, clinical considerations, and therapeutic approaches: the REPROGRAM consortium position paper. Front Immunol (2020) 11:1648.

14. Tisoncik JR, Korth MJ, Simmons CP, Farrar J, Martin TR, Katze MG. Into the eye of the cytokine storm. Microbiol Mol Biol Rev (2012) 76:16–32.

15. Wilson JG, Simpson LJ, Ferreira A-M, Rustagi A, Roque J, Asuni A, Ranganath T, Grant PM, Subramanian A, Rosenberg-Hasson Y. Cytokine profile in plasma of severe COVID-19 does not differ from ARDS and sepsis. JCI insight (2020) 5:

16. Lucas C, Wong P, Klein J, Castro TBR, Silva J, Sundaram M, Ellingson MK, Mao T, Oh JE, Israelow B. Longitudinal analyses reveal immunological misfiring in severe COVID-19. Nature (2020) 584:463–469.

17. Laing AG, Lorenc A, Del Molino Del Barrio I, Das A, Fish M, Monin L, Muñoz-Ruiz M, McKenzie DR, Hayday TS, Francos-Quijorna I. A dynamic COVID-19 immune signature includes associations with poor prognosis. Nat Med (2020) 26:1623–1635.

18. Silvin A, Chapuis N, Dunsmore G, Goubet A-G, Dubuisson A, Derosa L, Almire C, Hénon C, Kosmider O, Droin N. Elevated calprotectin and abnormal myeloid cell subsets discriminate severe from mild COVID-19. Cell (2020) 182:1401–1418.

19. Galani I-E, Rovina N, Lampropoulou V, Triantafyllia V, Manioudaki M, Pavlos E, Koukaki E, Fragkou PC, Panou V, Rapti V. Untuned antiviral immunity in COVID-19 revealed by temporal type I/III interferon patterns and flu comparison. Nat Immunol (2021) 22:32–40.

20. Ravindra NG, Alfajaro MM, Gasque V, Huston NC, Wan H, Szigeti-Buck K, Yasumoto Y, Greaney AM, Habet V, Chow RD. Single-cell longitudinal analysis of SARS-CoV-2 infection in human airway epithelium identifies target cells, alterations in gene expression, and cell state changes. PLoS Biol (2021) 19:e3001143.

21. Ziegler CGK, Miao VN, Owings AH, Navia AW, Tang Y, Bromley JD, Lotfy P, Sloan M, Laird H, Williams HB, et al. Impaired local intrinsic immunity to SARS-CoV-2 infection in severe COVID-19. Cell (2021) 184:4713–4733.e22. doi: https://doi.org/10.1016/j.cell.2021.07.023

22. Wang J, Li Q, Yin Y, Zhang Y, Cao Y, Lin X, Huang L, Hoffmann D, Lu M, Qiu Y. Excessive neutrophils and neutrophil extracellular traps in COVID-19. Front Immunol (2020) 11:2063.

23. Meizlish ML, Pine AB, Bishai JD, Goshua G, Nadelmann ER, Simonov M, Chang C-H, Zhang H, Shallow M, Bahel P. A neutrophil activation signature predicts critical illness and mortality in COVID-19. Blood Adv (2021) 5:1164–1177.

24. Zheng H-Y, Zhang M, Yang C-X, Zhang N, Wang X-C, Yang X-P, Dong X-Q, Zheng Y-T. Elevated exhaustion levels and reduced functional diversity of T cells in peripheral blood may predict severe progression in COVID-19 patients. Cell Mol Immunol (2020) 17:541–543.

25. Diao B, Wang C, Tan Y, Chen X, Liu Y, Ning L, Chen L, Li M, Liu Y, Wang G. Reduction and functional exhaustion of T cells in patients with coronavirus disease 2019 (COVID-19). Front Immunol (2020)827.

26. Sumon TA, Hussain MA, Hasan MT, Hasan M, Jang WJ, Bhuiya EH, Chowdhury AAM, Sharifuzzaman SM, Brown CL, Kwon H-J. A revisit to the research updates of drugs, vaccines, and bioinformatics approaches in combating COVID-19 pandemic. Front Mol Biosci (2021) 7:585899.

27. Panigrahy D, Gilligan MM, Huang S, Gartung A, Cortés-Puch I, Sime PJ, Phipps RP, Serhan CN, Hammock BD. Inflammation resolution: a dual-pronged approach to averting cytokine storms in COVID-19? Cancer Metastasis Rev (2020) 39:337–340.

28. Xu X, Han M, Li T, Sun W, Wang D, Fu B, Zhou Y, Zheng X, Yang Y, Li X. Effective treatment of severe COVID-19 patients with tocilizumab. Proc Natl Acad Sci (2020) 117:10970–10975.

29. Sungnak W, Huang N, Bécavin C, Berg M, Queen R, Litvinukova M, Talavera-López C, Maatz H, Reichart D, Sampaziotis F. SARS-CoV-2 entry factors are highly expressed in nasal epithelial cells together with innate immune genes. Nat Med (2020) 26:681–687.

30. Lukassen S, Chua RL, Trefzer T, Kahn NC, Schneider MA, Muley T, Winter H, Meister M, Veith C, Boots AW, et al. SARS-CoV-2 receptor ACE2 and TMPRSS2 are primarily expressed in bronchial transient secretory cells. EMBO J (2020) 39:e105114. doi: https://doi.org/10.15252/embj.20105114

31. Wilk AJ, Rustagi A, Zhao NQ, Roque J, Martínez-Colón GJ, McKechnie JL, Ivison GT, Ranganath T, Vergara R, Hollis T, et al. A single-cell atlas of the peripheral immune response in patients with severe COVID-19. Nat Med (2020) 26:1070–1076. doi: 10.1038/s41591-020-0944-y

32. Wilk AJ, Lee MJ, Wei B, Parks B, Pi R, Martínez-Colón GJ, Ranganath T, Zhao NQ, Taylor S, Becker W, et al. Multi-omic profiling reveals widespread dysregulation of innate immunity and hematopoiesis in COVID-19. J Exp Med (2021) 218:e20210582. doi: 10.1084/jem.20210582

33. Ren X, Wen W, Fan X, Hou W, Su B, Cai P, Li J, Liu Y, Tang F, Zhang F, et al. COVID-19 immune features revealed by a large-scale single-cell transcriptome atlas. Cell (2021) 184:1895–1913.e19. doi: https://doi.org/10.1016/j.cell.2021.01.053

34. Hao Y, Hao S, Andersen-Nissen E, Mauck III WM, Zheng S, Butler A, Lee MJ, Wilk AJ, Darby C, Zager M, et al. Integrated analysis of multimodal single-cell data. Cell (2021) 184:3573–3587.e29. doi: 10.1016/j.cell.2021.04.048

35. Aibar S, González-Blas CB, Moerman T, Huynh-Thu VA, Imrichova H, Hulselmans G, Rambow F, Marine J-C, Geurts P, Aerts J, et al. SCENIC: single-cell regulatory network inference and clustering. Nat Methods (2017) 14:1083–1086. doi: 10.1038/nmeth.4463

36. Shannon P, Markiel A, Ozier O, Baliga NS, Wang JT, Ramage D, Amin N, Schwikowski B, Ideker T. Cytoscape: A Software Environment for Integrated Models of Biomolecular Interaction Networks. Genome Res (2003) 13:2498–2504. doi: 10.1101/gr.1239303

37. Wu T, Hu E, Xu S, Chen M, Guo P, Dai Z, Feng T, Zhou L, Tang W, Zhan LI. clusterProfiler 4.0: A universal enrichment tool for interpreting omics data. Innov (2021) 2:100141.

38. Jin S, Guerrero-Juarez CF, Zhang L, Chang I, Ramos R, Kuan C-H, Myung P, Plikus M V, Nie Q. Inference and analysis of cell-cell communication using CellChat. Nat Commun (2021) 12:1088. doi: 10.1038/s41467-021-21246-9

39. Overmyer KA, Shishkova E, Miller IJ, Balnis J, Bernstein MN, Peters-Clarke TM, Meyer JG, Quan Q, Muehlbauer LK, Trujillo EA. Large-scale multi-omic analysis of COVID-19 severity. Cell Syst (2021) 12:23–40.

40. Chams N, Chams S, Badran R, Shams A, Araji A, Raad M, Mukhopadhyay S, Stroberg E, Duval EJ, Barton LM. COVID-19: a multidisciplinary review. Front public Heal (2020) 8:383.

41. Bruchez A, Sha K, Johnson J, Chen L, Stefani C, McConnell H, Gaucherand L, Prins R, Matreyek KA, Hume AJ. MHC class II transactivator CIITA induces cell resistance to Ebola virus and SARS-like coronaviruses. Science (80-) (2020) 370:241–247.

42. Laouedj M, Tardif MR, Gil L, Raquil M-A, Lachhab A, Pelletier M, Tessier PA, Barabé F. S100A9 induces differentiation of acute myeloid leukemia cells through TLR4. Blood, J Am Soc Hematol (2017) 129:1980–1990.

43. Yu K, He J, Wu Y, Xie B, Liu X, Wei B, Zhou H, Lin B, Zuo Z, Wen W. Dysregulated adaptive immune response contributes to severe COVID-19. Cell Res (2020) 30:814–816.

44. De Jong SJ, Créquer A, Matos I, Hum D, Gunasekharan V, Lorenzo L, Jabot-Hanin F, Imahorn E, Arias AA, Vahidnezhad H. The human CIB1–EVER1–EVER2 complex governs keratinocyte-intrinsic immunity to β-papillomaviruses. J Exp Med (2018) 215:2289–2310.

45. Schmitt J, Benavente R, Hodzic D, Höög C, Stewart CL, Alsheimer M. Transmembrane protein Sun2 is involved in tethering mammalian meiotic telomeres to the nuclear envelope. Proc Natl Acad Sci (2007) 104:7426–7431.

46. Sun W-W, Jiao S, Sun L, Zhou Z, Jin X, Wang J-H. SUN2 modulates HIV-1 infection and latency through association with lamin A/C to maintain the repressive chromatin. MBio (2018) 9:e02408–17.

47. Saeedi-Boroujeni A, Mahmoudian-Sani M-R. Anti-inflammatory potential of Quercetin in COVID-19 treatment. J Inflamm (2021) 18:1–9.

48. Mahmoudian-Sani M-R, Boroujeni AS, Khodadadi A, Houshmandfar S, Gandomkari ST, Alghasi A. Resveratrol; an inflammasome inhibitor and a potential therapy for severe cases of COVID-19. Immunopathol Persa (2021) 8:e9–e9.

49. Jones S, Rappoport JZ. Interdependent epidermal growth factor receptor signalling and trafficking. Int J Biochem Cell Biol (2014) 51:23–28. doi: https://doi.org/10.1016/j.biocel.2014.03.014

50. Venkataraman T, Frieman MB. The role of epidermal growth factor receptor (EGFR) signaling in SARS coronavirus-induced pulmonary fibrosis. Antiviral Res (2017) 143:142–150.

51. Ramírez-Martínez G, Jiménez-Álvarez LA, Cruz-Lagunas A, Ignacio-Cortés S, Gómez-García IA, Rodríguez-Reyna TS, Choreño-Parra JA, Zúñiga J. Possible Role of Matrix Metalloproteinases and TGF-β in COVID-19 Severity and Sequelae. J Interf Cytokine Res (2022)

52. Fernandez-Patron C, Hardy E. Matrix Metalloproteinases in Health and Disease in the Times of COVID-19. Biomolecules (2022) 12:692.

53. Carolina D, Couto AES, Campos LCB, Vasconcelos TF, Michelon-Barbosa J, Corsi CAC, Mestriner F, Petroski-Moraes BC, Garbellini-Diab MJ, Couto DMS. MMP-2 and MMP-9 levels in plasma are altered and associated with mortality in COVID-19 patients. Biomed Pharmacother (2021) 142:112067.

54. Zhang J-Y, Wang X-M, Xing X, Xu Z, Zhang C, Song J-W, Fan X, Xia P, Fu J-L, Wang S-Y. Single-cell landscape of immunological responses in patients with COVID-19. Nat Immunol (2020) 21:1107–1118.

55. Agresti N, Lalezari JP, Amodeo PP, Mody K, Mosher SF, Seethamraju H, Kelly SA, Pourhassan NZ, Sudduth CD, Bovinet C, et al. Disruption of CCR5 signaling to treat COVID-19-associated cytokine storm: Case series of four critically ill patients treated with leronlimab. J Transl Autoimmun (2021) 4:100083. doi: https://doi.org/10.1016/j.jtauto.2021.100083

